# Hearing the voices of Australian healthcare workers during the COVID-19 pandemic

**DOI:** 10.1101/2020.09.25.20197061

**Authors:** MR Ananda-Rajah, BG Veness, D Berkovic, C Parker, G Kelly, D Ayton

## Abstract

**Background:** The statistics of healthcare worker (HCW) COVID-19 infections do not convey the lived experience of HCWs during the pandemic. This study explores the working conditions and issues faced by Australian HCWs.

**Methods:** Qualitative analysis of free-text responses from Australian HCWs from 3 August to 5 August 2020 from an open letter calling for better respiratory protection for HCWs, transparent reporting of HCW COVID-19 infections and diversity in national infection control policy development. The open letter was sent to an email list of 23,000 HCWs from a previous campaign and promoted on social media.

**Results:** Among 2,733 HCWs who signed the open letter during the study period, 407 free-text responses were analysed. Doctors and nurses accounted for 58% and 35% of respondents, respectively. Most respondents came from Victoria (48%); New South Wales (18%); Queensland (12%) or Western Australia (12%). Dominant themes included concerns about: work health and safety standards; guidelines on respiratory protection including the omission of fit-testing of P2/N95 respirators; deficiencies in the availability, quality, appropriateness and training of personal protective equipment; a top-down workplace culture that enabled bullying in response to concerns about safety that culminated a loss of trust in leadership, self-reported COVID-19 infections in some respondents and moral injury.

**Conclusion:** Occupational moral injury in HCWs is the consequence of lapses in leadership at policy-making and organisational levels that have violated the normative expectations of HCWs. The challenge for healthcare leaders is to address workplace culture, consultation and engagement with HCWs in order to prevent this hidden pandemic from spreading throughout the health system.

## Introduction

Compared to many developed countries, Australia was relatively spared the impacts of COVID-19. Total case numbers were 7,767 until 30 June 2020 but increased when Victoria, a state with a population of >6 million, experienced a resurgence in early July 2020 accounting for 19,574 (75%) of a total of 26,207 infections by 6 September 2020 (1). This resurgence was associated with a sharp increase in healthcare worker (HCW) infections in Victoria (national data unavailable) from 388 on 16 July (2) to 3,206 by 3 September with one death reported in April 2020 (3).

There are parallels between the Australian and international experience with HCWs disproportionately infected with COVID-19 compared to the general community. By 16 September 2020, an estimated three million HCWs globally had been infected (4) and as of 16 July 2020 over 3000 had died (5). The prevalence of COVID-19 in HCWs from the United States and the United Kingdom is 2,747 cases per 100,000 HCWs, compared with 242 cases per 100,000 people in the general community (6). At least 1,000 HCWs have died in the USA (7) and 600 in the UK (5), with HCWs from non-Caucasian backgrounds comprising nearly two-thirds of deaths (8). While many jurisdictions have struggled to prevent HCW infections, some have effectively protected their HCWs, with China (9), Singapore (10) and Taiwan (11) the notable exceptions.

Statistics however, do not convey the lived experience of HCWs. In Australia, the media and professional societies have highlighted some of the issues faced by HCWs, who have reported workplace bullying (12), concerns about personal protective equipment (PPE) (13) and mental health morbidity (14). HCWs have turned to the media to help broadcast their concerns (15) in response to a lack of progress using conventional channels. The aim of this analysis was to give voice to HCWs about the challenges they have faced during the COVID-19 pandemic.

## Methods

### Data collection and analysis

On 2 August 2020, a letter signed by 23 doctors was sent to the Australian Minister for Health. The letter was shared via email to 23,000 HCWs who had responded to earlier campaigns, with an invitation to add their name in support. It garnered over 2,700 signatures by the morning of 5 August 2020. An optional question was included for respondents to share concerns about their occupational safety. Experienced health services researchers (DA, CP, DB), who were not involved in development of the open letter, coded the responses. Twenty responses were initially coded to create a coding framework. The primary codes were individual, organisational and system-level factors with barriers and enablers identified for each category. Approximately 130 responses were coded by each investigator, with coding disagreement resolved through discussion and consensus. Of note, HCWs are continuously responding to the open letter; this manuscript provides an interim analysis, particularly pertinent given increasing HCW COVID-19 infections. Ethics approval was received from Monash University (Project ID 26132).

### Results

The open letter was signed by 2,733 HCWs with 407 free-text contributions. The majority (48%) of respondents were from Victoria (VIC); followed by New South Wales (NSW) (18%); and similar representation from Queensland (QLD) and Western Australia (WA) (12%). Doctors and nurses accounted for 93% of responses, with remaining respondents from paramedicine, allied health, and clerical backgrounds (Table 1).

**Table 1.**
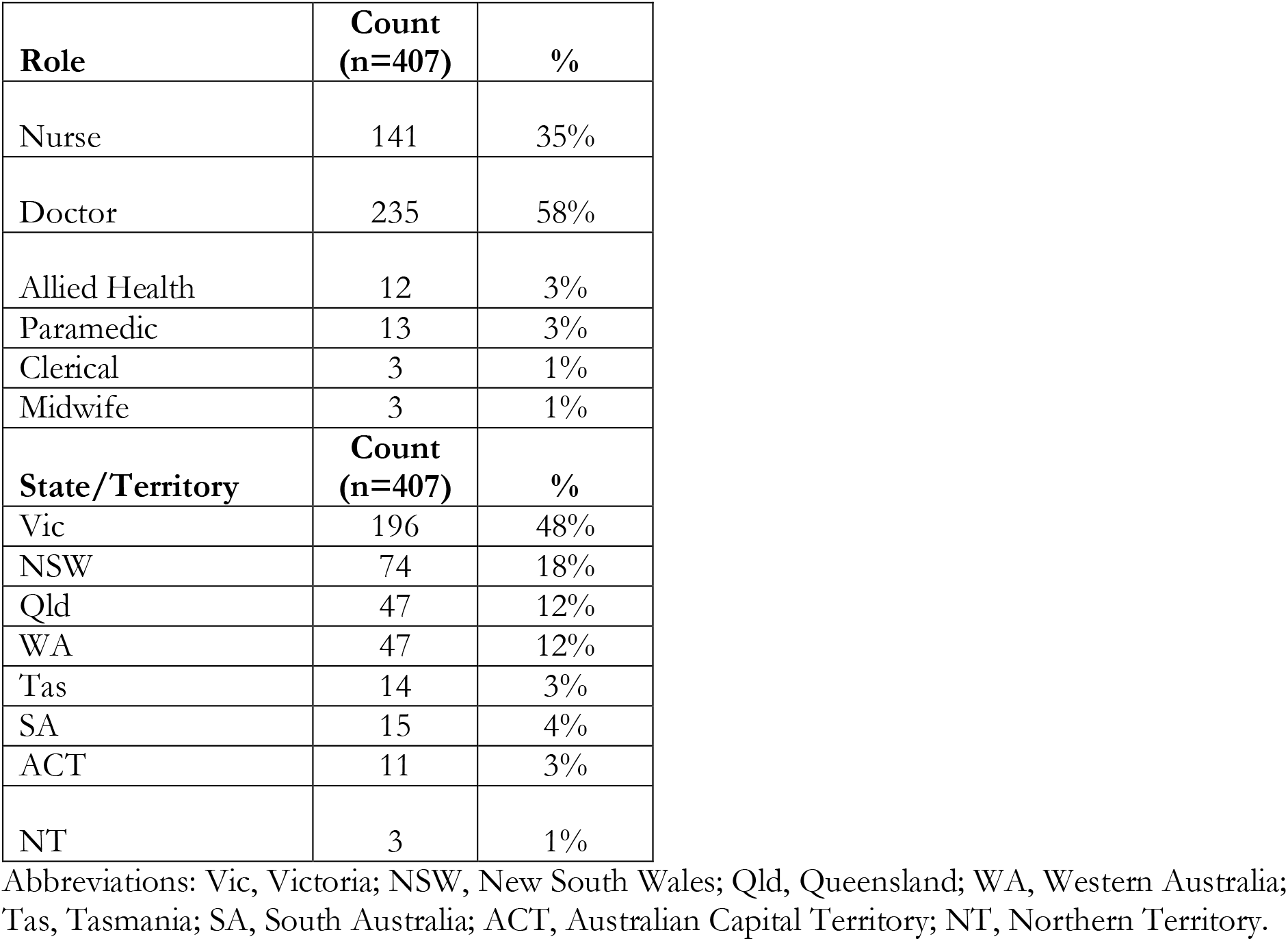
Demographic characteristics of healthcare worker respondents

| **Role** | **Count**<br>**(n=407)** | **%** |
| --- | --- | --- |
| Nurse | 141 | 35% |
| Doctor | 235 | 58% |
| Allied Health | 12 | 3% |
| Paramedic | 13 | 3% |
| Clerical | 3 | 1% |
| Midwife | 3 | 1% |
| **State/Territory** | **Count**<br>**(n=407)** | **%** |
| Vic | 196 | 48% |
| NSW | 74 | 18% |
| Qld | 47 | 12% |
| WA | 47 | 12% |
| Tas | 14 | 3% |
| SA | 15 | 4% |
| ACT | 11 | 3% |
| NT | 3 | 1% |

Abbreviations: Vic, Victoria; NSW, New South Wales; Qld, Queensland; WA, Western Australia; Tas, Tasmania; SA, South Australia; ACT, Australian Capital Territory; NT, Northern Territory.

The themes below represent the spectrum of issues concerning HCWs. To convey their voices, results have been presented in a narrative style below. Supplementary Table 1 provides HCW quotes mapped to the themes below.

“*We don’t want to be heroes. We just want protection in line with OHS standards* (GP, Victoria)” – Policies/guidelines and work health and safety obligations

HCWs felt that they deserve *“the same occupational health and safety demanded in any other industry”* (Anaesthetist, VIC) and that “*with community transmission rates climbing, subpar PPE is just unacceptable*.*”* (Trainee doctor, Vic). HCWs highlighted the mining and asbestos removal industry where “*no worker can work without the appropriate respirator”* (Nurse, WA). HCWs perceived guidelines derived from the World Health Organization to be “*woefully inadequate…*[*with*] *studies showing the virus to be airborne since March, whereas WHO only got on board in July*.*”* (GP, QLD). This WHO position gave “*managers and hospital executives an excuse to lower the standard of PPE to a surgical mask, face shield, and apron*” (Nurse, WA) with HCWs noting “*very poor leadership from executive*” (Doctor, VIC). Many perceived that guidelines were “*dictated by resources and not staff safety*” (Nurse, QLD) and that Australia has “*waited until our own staff got sick and intubated before we very gradually changed the PPE quality supplied to staff in Australia*.*”* (Doctor, VIC).

HCWs decried that “*staff becoming sick is unacceptable*” (Nurse, TAS) and “*having any fellow HCWs at the same health network with confirmed COVID is far too many”* (Doctor, VIC). They asked, “s*ince when was it acceptable for miners to die from preventable OHS accidents? Never - so don’t make it acceptable for HCWs”* (Doctor, VIC) noting that “*We give everything for our patients, but we don’t expect to have to give our lives”* (Nurse, TAS).

The approach to occupational safety was, *“reactive not proactive as numbers began to grow”* (Nurse, VIC). Instead of introducing “*PPE appropriate for airborne transmission, and scale back if required”* (Doctor, VIC), policies dismissed precaution and were “*dragging their heels … Adopting the cheapest PPE instead of the most suitable*.*”* (Paramedic, VIC). Respondents drew a comparison to SARS, where “*respiratory protection was upgraded well before the evidence-based studies regarding respiratory protection ‘proved’ that aerosol spread was occurring which saved hundreds of HCW lives”* (Physician, TAS).

Opportunities had been squandered given, “*Australia had so much time to learn from the experiences overseas”* (Nurse, TAS). Instead we “*lost valuable time to prepare for this pandemic due to government and department effective obstructiveness”* and did not consult broadly enough, “*occupational medicine doctors should have been involved in expert groups from start to prevent workplace outbreaks*.*”* (Occupational medicine physician, WA). HCWs recognised the threat to health system functionality where asymptomatic patients who had “*moved across many areas of the hospital”* could cause an outbreak “*affecting hospital flow and productivity*.*”* (Midwife, VIC)

“*Protect us PROPERLY so we can protect you!”* (Nurse, Western Australia)- Lack of access to PPE, issues with quality and appropriateness

Despite HCWs being “*essential*”, a lack of or limited supply of PPE was reported across Australia. Respondents emphasised that they *“need PPE in our practice consult rooms, not in a ‘National Stockpile’ thousands of kilometres away!”* (Doctor, QLD). The lack of availability of PPE had led to rationing, with PPE being “*locked in cupboards”* (Doctor, NSW) and respondents being *“directed to store surgical masks in a plastic bag and reuse them for the same patient on multiple occasions”* (Doctor, NSW). PPE had quality issues with “*gowns that tear easily, surgical mask quality is often poor”* (Nurse, WA), masks that *“often require a tremendous amount of* “*MacGuyver-ing”* but still *“slid down, exposing my nose”* (Trainee doctor, VIC), “*poorly fitting ear loop ones”* which affected critical manoeuvres where “*several colleagues had to remove eye protection as they kept fogging up*.*”* (Surgeon, NSW).

HCWs were angered by a lack of internal support: “*My hospital’s Chief Medical Officer has been silent on this issue, even when it was raised in a public forum*.*”* (Trainee doctor, VIC), citing that the inadequacy of PPE “*runs contrary to OH&S regulations in every other facet of working life”* (Trainee doctor, QLD). The lack of representation in policy-making meant that, “*General Practice is under-represented in the decision-making process and the allocation of PPE, despite our high profile in the early presentations”*.*”* (Nurse, WA). Consequently, HCWs are “*purchasing their own masks, at extreme cost*” (Nurse, SA) including non-approved items such as “*an elastomeric P3 filter mask but I would be disciplined if I wore it to work despite it likely being superior to a non-fit tested N95/P2 respirator*.*”* (Doctor, VIC).

*“We were horrified by the mismatch in different people’s faces to various brands of P2/N95”* (*GP, Tasmania*)*-* Fit-testing of respirators and PPE training

The provision of respirators is not a one-size-fits-all matter. Respiratory protection is only effective if HCWs “*have quantitative Fit Testing*” (Anaesthetist, VIC) “*to ensure that they provide an adequate seal. Without proper fitting, these masks provide no additional protection over and above a standard surgical mask*.*”* (Anaesthetist, NSW). One surgeon recounted how they “*had to apply strips of Micropore tape around the mask to achieve an adequate seal. … It beggars belief that HCWs are expected to jury-rig N95 masks to achieve a life-protecting air-tight seal”* (Surgeon, WA).

In-house fit-testing was occurring, with alarming results, where “*around 40% of women failed a quantitative fit test on the disposable P2/N95 masks”* (Anaesthetist, NSW), noting that *“women and non-Caucasian faces appear at greatest risk of failure. They also appear to be disproportionately the HCWs exposed and dying in the UK*.*”* (Anaesthetist, QLD). Arranging fit-testing was not easy, with HCWs having to *“fight to even be fit-tested for N95 masks”*, and having to *“to pay for that fit-testing myself before the hospital relented and tested the rest of the department*.*”* (Doctor, TAS). HCWs were bewildered as to *“why certain industry sectors make it mandatory for their workers to have such rigorous testing performed and others such as healthcare, where lives matter just as much, do not seem to be implementing this”* (Anaesthetist, VIC). *“Supply issues”* was the reason cited for organisational resistance to fit-testing even when there were “*multiple HCW infections despite adhering to the current policy for PPE”* (ED doctor, VIC).

Reusable respirators such as powered air-purifying respirators (PAPRs) were seen as viable alternatives to disposable respirators “*as it provides superior protection in those who cannot achieve adequate seal with N95 or P2 masks”* (Anaesthetist, WA) and had a proven track record in safety during SARS: “*I owe my life to the wearing of the 3M Jupiter Hood System with ventilatory filters…issued by the hospital administration*.*”* (Physician, WA). However, there was resistance to reusables even when “*none of the N95 masks fit my face and my hospital won’t let me wear my own 3M respirator, so how do I minimise my risk”?* (Anaesthetist, NSW).

Fit for purpose PPE was important particularly for first responders where they “*do not function in a controlled environment”* and *“face wind, weather, manual handling of infected patients, difficult extrications, patient lifts, carries, and sit centimetres from patients in confined spaces”*. Gowns were prone *“to billowing in the wind, spreading droplets, contaminating faces and surfaces, as well as providing no protection to legs when bending, lifting, sitting”* (Paramedic WA). Proceduralists encountered PPE failures, “*on a patient who had suffered a cardiac arrest, the mask developed a major leak and fogged up the face shield which meant I was exposed to viral aerosol and had poor visibility*.*”* (Anaesthetist, VIC). Even anaesthetists, who are at high risk due to intubation responsibilities are being asked “*to use cheap N95 duckbill masks without formal fit testing… intubators’ face shields are fogging so badly that it is almost impossible to see. Full face Respirators must be made available*.*”* (Anaesthetist, VIC). Guideline variability resulted in mask confusion with the need “*to juggle between different masks based on the interventions we use to treat patients”* where *“we are not able to predict these decisions until we are already on scene”* (Paramedic, NSW).

In addition to fit testing, PPE training was required. *“Donning and doffing correctly, needs education and compliance monitoring! I’ve seen so many nurses do it incorrectly”* (Surgical Nurse, VIC). There were calls for *“a nationwide respiratory protection program incorporating the training, use and centralised cleaning of PAPRs and reusable elastomeric respirators for those who do not fit disposable P2/N95 masks*.*”* Without this level of oversight, “*Governments and health care organisations are currently failing in their duty of care to provide a safe working environment for HCWs*.*”* (Anaesthetist, NSW).

“*I have not been sleeping well for a long time. I am likely to be called up for active service on the COVID wards and this fear is playing on my mind*” (Physician, Victoria) -Fear for self and others bullying, victimisation and censure

HCWs described being bullied and victimised in their workplaces, being told they were *“not a team player”* (ED doctor, NSW) and “*face shaming tactics from colleagues”* (GP, WA), in a workplace that *“has actively tried to silence me”* (Allied Health, QLD) for expressing concerns about their safety. HCWs were “*threatened to be stood down for requesting an N95 mask”* (Paediatrician, NSW), having “*N95 masks taken out of our hands before going into see positive COVID patients*.*”* (Physician, VIC) and of being “*bullied by admin staff who don’t want us to ask for appropriate PPE, and lied to”* (Anaesthetist, QLD).

Senior managers had told staff that they *“need to toughen up”* (ED Doctor, QLD) while infection control nurses were *“chastising front-line ED nurses caring for suspected COVID-19 patients for wearing N95s, saying they should only use surgical masks”* (Trainee doctor, TAS). Requests for use of personal higher-grade PPE was discouraged by infectious diseases experts as it may *“‘set a precedent and that the optics of not following the* [*Department of Health and Human Services*] *guidelines were not good*.*”* (Nurse, VIC). Infection control were “*very poor in backing up the HCWs”* and preferred to *“toe the executive line rather than evidence-based practice*.*”* (ED physician, NSW), creating *“a very bad taste in our department”* (ED physician, NSW). HCWs were angry that infectious diseases experts “*had determined N95 masks were only needed for aerosol generating procedures in COVID19 patients”* and who expressed “*hubris when they were asked about HCW infections”* implying *“that the high rate of HCW infections and deaths was due to incorrect donning and doffing rather than the grade of the PPE itself”* (Physician, VIC).

Concerns about workplace safety had “*caused increased anxiety,”* (Nurse, WA), leading to withdrawal from family noting that being *“away from my family and my pets is hard emotionally, but I do this so I can serve my community and protect my family”* (Doctor, WA). HCWs were “*mentally exhausted stressing about the possibility of bringing home the virus”* (Trainee Doctor, WA), with *“growing burnout … We are losing sleep at unprecedented levels due to worry*.*”* (GP, NSW). They felt “*threatened coming to work and seeing COVID suspected patients with inadequate PPE”* (Doctor, WA).

*“I was denied N95 masks, informing me they are “unnecessary” when caring for COVID19 positive patients. I am now COVID19 positive, and won’t see my baby for weeks. All because of trying to care for people, when no one cared for me*.*”* (*Nurse, VIC*)*-* Self-disclosed HCWs infections

HCWs stated that COVID-19 infections were “*almost certainly contracted in my workplace*.*”* (Doctor, COVID clinic, VIC) as “*evidenced in 10% of cases being HCWs. Rates in the community are far lower than 10%. HCWs are acquiring this infection in their places of employment*.*”* (Anaesthetist, VIC). Many felt that they contracted COVID-19 despite *“wearing the correct PPE, and yet it still wasn’t safe enough”* (Nurse, VIC). PPE guidelines were ineffective as “*three nurses on our ward tested positive to COVID-19, only surgical masks were provided to nurses when caring for suspected positive patients*.*”* (Nurse, VIC). Similar stories were evident for community-based HCWs: *“my colleague looked after two COVID patients in their home wearing the current PPE and is now at home isolating as he is COVID positive*. (Physician, VIC). “*The risk is real and it is too great not to act”* said an infected ED Doctor (VIC) especially with a “*close friend in ICU after contracting COVID working in ED”* (GP, VIC).

“*We have the knowledge and skills required to sort this out. The bureaucracy is getting in the way of safety*.*”* (Anaesthetist, NSW)*-* Organisational barriers, poor consultation, loss of trust in leadership

Underlying the themes was a loss of trust in leadership at service, state and national levels. “*We use our judgement and analyse situations as part of our daily work. Do not lie to us and gaslight us when we assess the situation to be unacceptably risky”* (ED doctor, WA). Throughout the pandemic “*there is one thing which is constant and consistent, which is no transparency and no honesty reflected in the leadership’s policies”* (ED doctor, NSW). *“We have had very poor leadership from executive, they have cut corners with our safety at every turn”* (Physician, VIC), such that *“I have lost faith in leadership that sees risk to my safety at work as acceptable*.*”* (Trainee doctor, VIC). Directives were top-down with little consultation with HCWs because, “*Hospital admin only take advice from the health department despite requests and recommendations by local physicians*” (Physician, NSW). Requests for extra precautions were “*not responded* [*to*] *at all despite repeated requests and lobbying”* (Physician, VIC). HCWs found *“it hard to trust leadership”* which *“makes already low morale, even lower”* (Physician, VIC) and were “*seriously thinking of leaving nursing due to the lack of leadership, support and respect within the system*.*”* (Nurse, NSW). The lack of precaution was short sighted with HCW wondering *“what happens when we’ve infected all our staff and don’t have adequate staffing levels to provide care”* (Anaesthetist, NSW).

## Discussion

This study is a clarion call for organisational leaders, managers and policy makers to acknowledge and address the barriers to work, health and safety that have violated the normative expectations of healthcare workers. The HCW respondents in this study expect to feel safe at work but find themselves in the opposite situation due in a large part, to leadership not meeting their expectations at national, state and local service levels. Themes inductively identified from the data spanned the system, organisational and personal impacts of COVID-19 with lapses in leadership cross cutting all of them. Policies on respiratory protection were perceived to have disregarded the precautionary principle that is implicit within work health and safety policy (16). Issues regarding access to PPE, its appropriateness and quality; a lack of respirator fit-testing within a respiratory protection program; and a command-and-control culture in the workplace that suppressed respondent concerns, was felt to have contributed to self-reported workplace acquisitions of COVID-19. Respondents were critical of leadership, at all jurisdictional levels, which has resulted in a loss of trust that threatens to endure for years to come. The moral injury to HCWs would appear to be the “hidden pandemic”, resulting in emerging mental health issues including anxiety, sleeplessness, withdrawal, resentment and anger.

Respondents reported being in the untenable position of needing to deliver patient care while facing unacceptably high risks to themselves in a workplace failing to protect them (17, 18). The list of injurious events started with concerns about national guidelines on respiratory protection endorsing the surgical mask for routine care of COVID-19 patients rather than fit-tested P2/N95 respirators or above. To respondents, academic arguments of disease transmission referencing the aerosol versus droplet dichotomy (19) were resolved early in the pandemic. Instead, they felt forced to follow policies at odds with their own assessment of personal risk. Raising legitimate concerns appeared to have invited bullying, victimisation and censure within their organisations. Respondents cited a disregard of work health and safety obligations bordering on hubris by organisational leaders and infection control/infectious diseases experts, who were unfortunately perceived as being more intent on enforcing guidelines rather than meaningful engagement with frontline staff. As a result, several HCWs took matters into their own hands, organising in-house fit-testing or the purchase of reusable respirators.

During this pandemic, moral injury has referred to challenging decisions involving patient care that conflict with provider values (20). The concept originated in the military (21) and was extended by Litz *et al*. to “perpetrating, failing to prevent, bearing witness to, or learning about acts that transgress deeply held moral beliefs and expectations” (22). A complementary variant has emerged from the social sciences in reference to healthcare where “it arises from sources that include injustice, cruelty, status degradation and profound breaches of moral expectations” (23). It is not a mental illness, but rather a violation of an individual’s moral or ethical code that results in psychological distress (20). The voices of the respondents clearly point to an established moral injury where they feel like they are being treated as *“expendable”* rather than essential to the pandemic response.

Reversing occupational moral injury is not easy once established but this is the challenge healthcare leadership must rise to. Shale, in a seminal and timely commentary, presents a practical roadmap for moral repair (23). This framework prefaces each of the seven actions with acknowledgment: of the injured party as a moral equal, of shared norms, of testimony in a climate of safety, of the responsibility of leaders which is not equivalent to directing blame at them, of remediation to rectify the issues, of negative feelings and finally, of authentic acknowledgement of sorrow and regret that are not rehearsed apologies (23). The high degree of engagement required for reparation stands in contrast to the limited consultation experienced by many respondents on matters relating to their safety at work. Inadequate consultation has been a missed opportunity that also contravenes Australian Work Health and Safety legislation designed to address the power imbalance between management, who issue directives, and workers placed in the risk zone (16).

This study has several limitations. The purpose of the open letter was primarily to advocate for increased protection for HCWs, and thus the methodology was not designed to optimally probe HCW perspectives. The study may not be representative of all HCWs as the response rate was 12% at 48 hours and it did not capture all types of HCWs including a large sample of nurses, administrative or support staff. Occupational moral injury has not been well described in healthcare and deserves further research to better understand its root causes, preventative strategies and evidence based remediation. Response and non-response bias common to survey methodology may be operating and these findings require confirmation in studies from other jurisdictions. Finally, there was no ‘check’ to ensure that respondents were indeed HCWs, however the consistency of themes would argue otherwise.

Constructive engagement with HCWs that also addresses their moral injury presents the challenge for healthcare leaders during this pandemic and beyond. This process should be seen as an opportunity to harness the resourcefulness of HCWs that many respondents felt had been sidelined, with the wealth of governance and operational experience among healthcare leaders, policy makers and government in order to fast track solutions. Achieving shared goals necessarily starts with better consultation and a dose of courage in all actors. Not doing so risks the contagion described in this report spreading throughout Australia’s healthcare system with implications for other jurisdictions.

## Data Availability

The open letter is available: https://tinyurl.com/PPE4HCWs. Individual responses to the open letter will be made available pending ethics approval in an open source repository if the study is accepted by a peer review
journal.

